# How Often Do Patients Need Retreatment After Surgery for Subdural Hematoma? A Nationwide Readmission Database Analysis

**DOI:** 10.1101/2023.07.28.23293352

**Authors:** Pouya Nazari, Pedram Golnari, William Nathaniel Metcalf-Doetsch, Matthew B. Potts, Babak S. Jahromi

## Abstract

**Background and purpose:** The incidence of subdural hematoma (SDH) is increasing as the average age in the US rises, with reported high recurrence and reoperation rates. We therefore aimed to have a better understanding of retreatment following surgical evacuation of SDH in real-world practice.

**Methods:** Data were extracted from the latest available Nationwide Readmissions Database (2016-2019). Adult patients diagnosed with SDH who had burr-hole or craniotomy were included in the study.

**Results:** Retreatment rates were relatively low in both cohorts, with patients in the burr-hole cohort having a slightly higher rate of retreatment compared to the craniotomy cohort (8.4% vs 6.6%, p<0.001). The majority (>95%) of retreatments occurred within 90 days of initial treatment, and further follow up did not demonstrably increase retreatment on Kaplan Meier analysis. Retreatment rates remained consistent during the four-year study period in both burr-hole (7.2%-10.4%), and craniotomy cohorts (6.4%-6.8%).

**Conclusions:** Analysis of a large national database of unselected patients shows retreatment rates after surgery for SDH are lower than suggested by prior studies. Almost all retreatments occur within 90 days after initial treatment, which may have implications for length of follow up for such patients.

## Introduction

Subdural hematoma (SDH) is one of the most common types of intracranial hemorrhage, with more than 90,000 hospitalizations in the US per year. ^1,2^ The frequency of hospitalization for SDH has been increasing due to an aging population, and SDH is projected to be the most common cranial neurosurgical condition in the US by the year 2030. ^3,4^ Classically, patients with primary or recurrent symptomatic SDH are treated with surgical evacuation using burr-hole or craniotomy. ^5^ However, these surgical interventions are reportedly associated with high recurrence and reoperation rates of up to 30%,^5-7^ leading to the increasing popularity of middle meningeal artery (MMA) embolization as a treatment option for chronic SDH. ^8^ Nevertheless retreatment rates in patients with SDH have not been well described on a national scale. We therefore utilized the Nationwide Readmission Database (NRD) to characterize the rate of 6-month retreatment in SDH patients after burr-hole or craniotomy in the largest real-world cohort analyzed to date.

## Methods

### Database

Annual Nationwide Readmission Database (NRD) datasets (2016-2019) were obtained from the Healthcare Cost and Utilization Project (HCUP) Central Distributor (Rockville, Maryland, USA). To produce national estimates, discharge weights provided by the HCUP website were used. HCUP databases lack unique patient identifiers, and therefore, are exempt from Institutional Review Board review and informed consent under HIPAA. ^9^

### Patient Selection, Definitions, and End Point Variables

International Classification of Diseases, Tenth Revision, Clinical Modification (ICD-10-CM) codes were used to define medical diagnoses and inpatient procedures (Supplementary Table 1). Adult patients diagnosed with SDH who had burr-hole or craniotomy were included in the study. As the NRD only identifies readmissions within a single calendar year, initial encounters were only selected from the first six calendar months, to ensure a subsequent six-month follow-up period was available for the entire cohort each year. ^10,11^ Throughout the study period, ≤3% of procedures (≤0.2% of all patients) involved MMA embolization during the initial admission. We therefore excluded such patients and focused only upon patients undergoing surgical evacuation at initial admission. Repeat surgical evacuation during the initial admission or any readmission was considered as retreatment. Relevant comorbidities as well as the severity of illness and risk of mortality were analyzed as described previously. ^10,11^ The All Patient Refined Diagnosis Related Groups (APR-DRG) Classification System was used to classify the severity of illness and risk of mortality.

### Statistical Analysis

Student’s t- and chi-squared tests were performed in between-group comparisons for continuous and categorical variables, respectively, using the software IBM SPSS Statistics for Windows, Version 26 (IBM Corp., Armonk, NY). A p-value level of <0.05 was defined as statistically significant. Data are presented as mean ± standard deviation (SD) and 95% confidence intervals (CI) are presented as brackets unless otherwise specified. Kaplan-Meier curves and log-rank test were applied to compare the time to retreatment after craniotomy and burr-hole with censoring at time of first retreatment or death.

## Results

Based on the above inclusion criteria, we identified a total of 30,838 SDH patients who were admitted and treated from 2016 to 2019. Among these admissions, 15,806 (51.3%) patients (mean age 69.4 years ± 15.0 [SD], 69.9% male) underwent craniotomy, and 15,032 (48.7%) patients (mean age 72.2 years ± 13.2 [SD], 70.8% male) had burr-hole. There was a significant difference in age distribution between craniotomy and burr-hole (p<0.001), with craniotomy seen more frequently in younger patients and burr-hole more often performed in older patients. Baseline demographics and characteristics of the study population are shown in Table 1.

**Table 1.** Comparison of baseline characteristics of SDH patients, 2016-2019.

| Factors | SDH (n=30838) |  | p-value |
| --- | --- | --- | --- |
|  | Craniotomy (n=15806) | Burr hole (n=15032) |  |
|  | No. of events (%) | No. of events (%) |  |
| Age Groups |  |  | < 0.001 |
| <65 years | 4895 (31.0%) | 3623 (24.1%) |  |
| 65-79 years | 6467 (40.9%) | 6434 (42.8%) |  |
| $\geq 80$ years | 4444 (28.1%) | 4975 (33.1%) | |
| Sex |  |  | 0.096 |
| Female | 4756 (30.1%) | 4393 (29.2%) |  |
| Male | 11050 (69.9%) | 10639 (70.8%) |  |
| APR-DRG Mortality Risk |  |  | < 0.001 |
| Minor | 3126 (19.8%) | 4159 (27.7%) |  |
| Moderate | 4131 (26.1%) | 4077 (27.1%) |  |
| Major | 4814 (30.5%) | 4636 (30.8%) |  |
| Extreme | 3734 (23.6%) | 2160 (14.4%) |  |
| APR-DRG Severity |  |  | < 0.001 |
| Minor | 5073 (32.1%) | 4296 (28.6%) |  |
| Moderate | 2936 (18.6%) | 4246 (28.2%) |  |
| Major | 3800 (24.0%) | 4209 (28.0%) |  |
| Extreme | 3996 (25.3%) | 2281 (15.2%) |  |
| Comorbidity* |  |  | < 0.001 |
| Score (Mean ± SD) | 3.21 ± 2.04 | 3.09 ± 2.03 | |

APR-DRG = All Patients Refined Diagnosis Related Groups.
\* Elixhauser comorbidity index score (min=0, max=29)

### Retreatment Rates

The total retreatment rate within the 6-month follow-up period was 7.5% [95% CI, 7.2% to 7.8%], with patients in the burr-hole cohort having a higher rate of retreatment compared to the craniotomy cohort (8.4% [95% CI, 8.0% to 8.9%] vs 6.6% [95% CI, 6.2% to 7.0%], p<0.001) (Figure 1A). Retreatment rates remained consistent during the study period in both burr-hole (7.2%-10.4%), and craniotomy cohorts (6.4%-6.8%) (Figure 1B).

**Figure 1.**
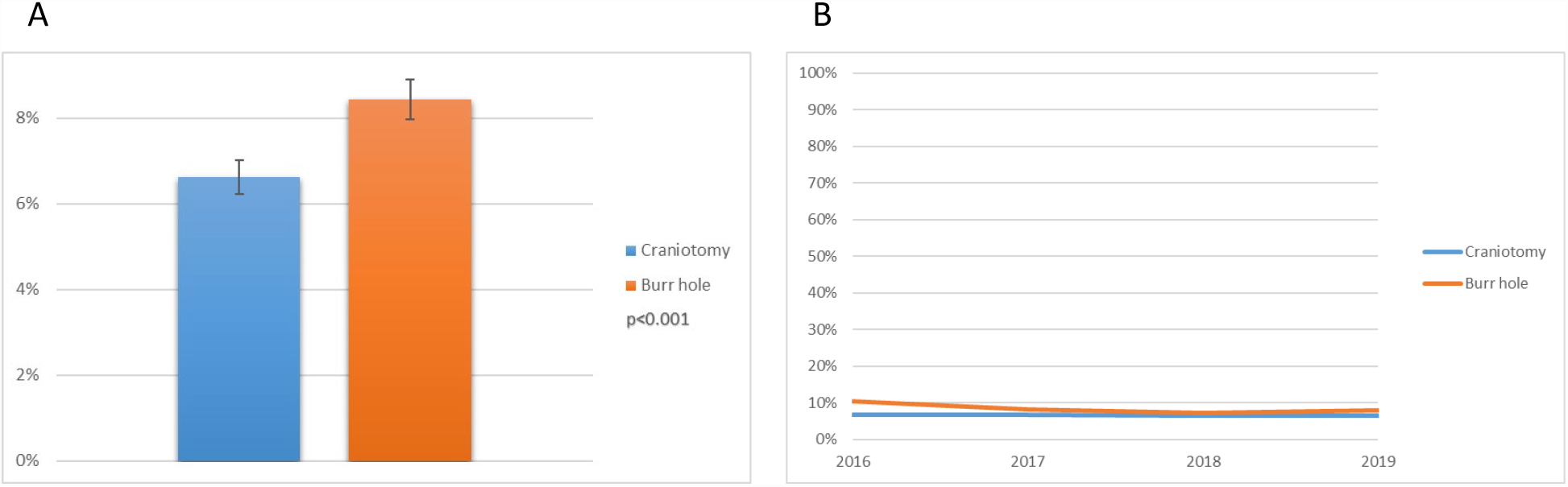
Overall (A) and annual (B) retreatment rates after craniotomy and burr hole in SDH patients (2016-2019).

Kaplan-Meier curves confirmed a higher rate of retreatment after burr-hole compared to craniotomy (Figure 2, log-rank: p<0.001). During follow up, 75.4% of all retreatment procedures occurred within 30 days after initial treatment, and 96.9% of all retreatment procedures occurred within the 90 days after initial treatment.

**Figure 2.**
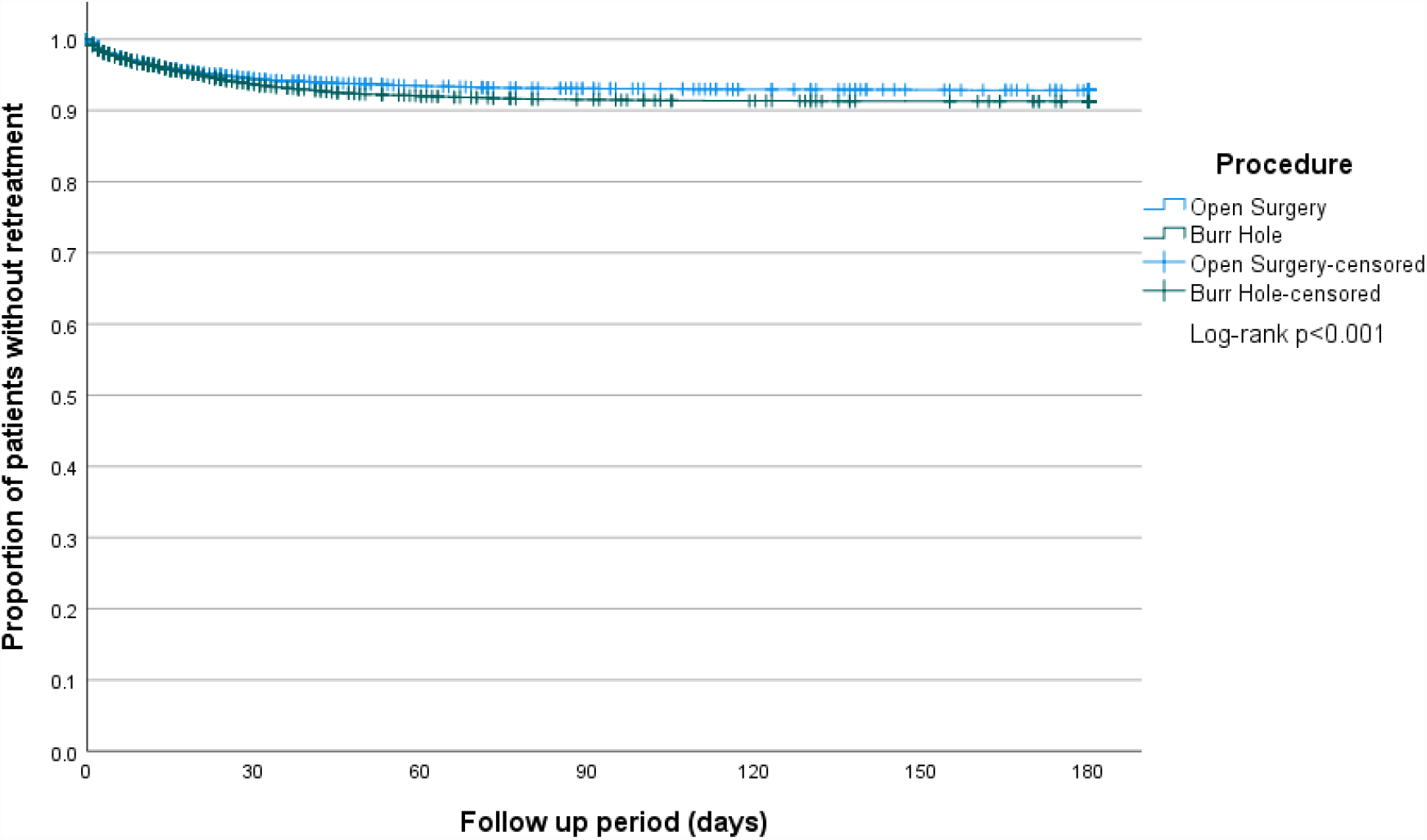
Kaplan-Meier graph showing the proportion of patients without retreatment after craniotomy or burr-hole.

## Discussion

The incidence rate of SDH is projected to increase progressively as the average age in US continues to rise. ^3,4^ An accurate understanding of retreatment rates in real-world practice is therefore important for all neurovascular practitioners. Using the latest available nationally representative NRD database, we found only a small proportion of patients (6.5-8.5%) required retreatment. This retreatment rate is substantially smaller than the frequently quoted high recurrence and reoperation rate “of up to 30%”,^5-7^ and is lower or within range of other previous retrospective studies and clinical trials examining single-center or smaller study populations (5.0%-27.5%; Table 2). ^6,12-21^ We believe our data is unique in comparison to these prior studies in that we used the largest available nationally representative database to provide a more representative estimate of real-world practice than randomized trials or registries and selected case series which are subject to publication bias. Our results also suggest that a 3-month follow-up period should suffice for most SDH patients, as more than 95% of retreatments occur within 90 days of initial treatment.

**Table 2.** Review of literature.

| Author | Year | Design | Patients (n) | Institution(s) | Retreatment (%) | Follow up |
| --- | --- | --- | --- | --- | --- | --- |
| *Burr hole* |  |  |  |  |  |  |
| Santarius et al.<sup>12</sup> | 2009 | RCT | 215 | Single | 16.7% | 6 months |
| Javadi et al.<sup>13</sup> | 2011 | RCT | 40 | Single | 5.0% | 6 months |
| Bellut et al.<sup>14</sup> | 2012 | Retrospective | 113 | Single | 9.7% | 3 months |
| Gernsback et al.<sup>15</sup> | 2016 | Retrospective | 215 | Single | 6.1% | ? |
| Sjåvik et al.<sup>16</sup> | 2017 | Retrospective | 1,260 | Multiple | 13.4% | 6 months |
| Soleman et al.<sup>17</sup> | 2019 | RCT | 220 | Single | 10.0% | 12 months |
| Jensen et al.<sup>18</sup> | 2021 | RCT | 420 | Multiple | 13.3% | 3 months |
| Amoo et al.<sup>19</sup> | 2021 | Retrospective | 131 | Single | 27.5% | 3 months |
| Duerinck et al.<sup>6</sup> | 2022 | RCT | 79 | Multiple | 7.6% | 6 months |
| Bartley et al.<sup>20</sup> | 2022 | RCT | 541 | Multiple | 10.2% | 6 months |
| *Open surgery* |  |  |  |  |  |  |
| Lukasiewicz et al.<sup>21*</sup> | 2016 | Retrospective | 746 | Multiple | 8% | 1 month |
| Amoo et al.<sup>19</sup> | 2021 | Retrospective | 11 | Single | 9.1% | 3 months |
| Duerinck et al.<sup>6</sup> | 2022 | RCT | 84 | Multiple | 13.1% | 6 months |

\*Included all SDH subtypes. All other papers examined only chronic SDH.

Recent studies have proposed MMA embolization as an emerging minimally invasive therapy which can be an alternative or adjunct to surgical evacuation for primary or recurrent SDH. ^22,23^ Since our data implies a lower retreatment rate for SDH than previously published, the target population for adjuvant MMA embolization may therefore be smaller than believed. This also has implications for clinical trials of adjuvant MMA embolization: assuming an 8% need for retreatment, a clinical trial of adjuvant MMA embolization would require approximately 1100 patients (550 in each arm) to show a 50% reduction in retreatment rate after surgery (this does not address primary MMA embolization instead of surgery for SDH).

An important limitation of the NRD is that ICD-10 codes may not accurately define acute or chronic SDH. However, assuming burr-hole procedures are generally performed only for chronic SDH, the latter still demonstrated low retreatment rates in our dataset. Additionally, since ICD-10 codes do not allow us to distinguish initial bilateral SDH treatment from unilateral SDH retreatment, our low retreatment rates may actually be an overestimation. Finally, other limitations to the use of the NRD include known caveats associated with analysis of retrospective administrative databases. ^24^

## Conclusions

Using a large representative national database, we show that retreatment rates after surgery for SDH are lower than those often quoted in literature, with most retreatments occurring within 3 months of initial treatment. These data have implications for management and follow-up of SDH in clinical practice, and additionally suggest rigorous randomized trials are necessary before routine incorporation of newer adjuncts such as MMA embolization aimed at reducing SDH recurrence and/or retreatment.

## Data Availability

The data that support the findings of this study are available on request from the Healthcare Cost and Utilization Project (HCUP).

https://hcup-us.ahrq.gov/

## References

1. Franko LR, Sheehan KM, Roark CD, Joseph JR, Burke JF, Rajajee V, Williamson CA. A propensity score analysis of the impact of surgical intervention on unexpected 30-day readmission following admission for subdural hematoma. Journal of neurosurgery. 2018;129:1008–1016. doi: 10.3171/2017.6.Jns17188

2. Frontera JA, Egorova N, Moskowitz AJ. National trend in prevalence, cost, and discharge disposition after subdural hematoma from 1998-2007. Crit Care Med. 2011;39:1619–1625. doi: 10.1097/CCM.0b013e3182186ed6

3. Balser D, Farooq S, Mehmood T, Reyes M, Samadani U. Actual and projected incidence rates for chronic subdural hematomas in United States Veterans Administration and civilian populations. Journal of neurosurgery. 2015;123:1209–1215. doi: 10.3171/2014.9.Jns141550

4. Neifert SN, Chaman EK, Hardigan T, Ladner TR, Feng R, Caridi JM, Kellner CP, Oermann EK. Increases in Subdural Hematoma with an Aging Population-the Future of American Cerebrovascular Disease. World neurosurgery. 2020;141:e166–e174. doi: 10.1016/j.wneu.2020.05.060

5. Catapano JS, Scherschinski L, Rumalla K, Srinivasan VM, Cole TS, Baranoski JF, Lawton MT, Jadhav AP, Ducruet AF, Albuquerque FC. Emergency Department Visits for Chronic Subdural Hematomas within 30 Days after Surgical Evacuation with and without Middle Meningeal Artery Embolization. AJNR American journal of neuroradiology. 2022. doi: 10.3174/ajnr.A7572

6. Duerinck J, Van Der Veken J, Schuind S, Van Calenbergh F, van Loon J, Du Four S, Debacker S, Costa E, Raftopoulos C, De Witte O, et al. Randomized Trial Comparing Burr Hole Craniostomy, Minicraniotomy, and Twist Drill Craniostomy for Treatment of Chronic Subdural Hematoma. Neurosurgery. 2022;91:304–311. doi: 10.1227/neu.0000000000001997

7. Ducruet AF, Grobelny BT, Zacharia BE, Hickman ZL, DeRosa PL, Andersen KN, Sussman E, Carpenter A, Connolly ES, Jr. The surgical management of chronic subdural hematoma. Neurosurg Rev. 2012;35:155–169; discussion 169. doi: 10.1007/s10143-011-0349-y

8. Rudy RF, Catapano JS, Jadhav AP, Albuquerque FC, Ducruet AF. Middle Meningeal Artery Embolization to Treat Chronic Subdural Hematoma. Stroke: Vascular and Interventional Neurology. 2023;3:e000490. doi: doi:10.1161/SVIN.122.000490

9. DUA Training-Accessible Version. Healthcare Cost and Utilization Project (HCUP). https://www.hcup-us.ahrq.gov/DUA/dua_508/DUA508version.jsp. Accessed May 01, 2020.

10. Nazari P, Golnari P, Ansari SA, Cantrell DR, Potts MB, Jahromi BS. Unplanned readmission after carotid stenting versus endarterectomy: analysis of the United States Nationwide Readmissions Database. Journal of neurointerventional surgery. 2023;15:242–247. doi: 10.1136/neurintsurg-2021-018523

11. Golnari P, Nazari P, Ansari SA, Hurley MC, Shaibani A, Potts MB, Jahromi BS. Endovascular Thrombectomy after Large-Vessel Ischemic Stroke: Utilization, Outcomes, and Readmissions across the United States. Radiology. 2021;299:179–189. doi: 10.1148/radiol.2021203082

12. Santarius T, Kirkpatrick PJ, Ganesan D, Chia HL, Jalloh I, Smielewski P, Richards HK, Marcus H, Parker RA, Price SJ, et al. Use of drains versus no drains after burr-hole evacuation of chronic subdural haematoma: a randomised controlled trial. Lancet (London, England). 2009;374:1067–1073. doi: 10.1016/s0140-6736(09)61115-6

13. Javadi A, Amirjamshidi A, Aran S, Hosseini SH. A randomized controlled trial comparing the outcome of burr-hole irrigation with and without drainage in the treatment of chronic subdural hematoma: a preliminary report. World neurosurgery. 2011;75:731–736; discussion 620-733. doi: 10.1016/j.wneu.2010.11.042

14. Bellut D, Woernle CM, Burkhardt JK, Kockro RA, Bertalanffy H, Krayenbühl N. Subdural drainage versus subperiosteal drainage in burr-hole trepanation for symptomatic chronic subdural hematomas. World neurosurgery. 2012;77:111–118. doi: 10.1016/j.wneu.2011.05.036

15. Gernsback J, Kolcun JP, Jagid J. To Drain or Two Drains: Recurrences in Chronic Subdural Hematomas. World neurosurgery. 2016;95:447–450. doi: 10.1016/j.wneu.2016.08.069

16. Sjåvik K, Bartek J, Jr., Sagberg LM, Henriksen ML, Gulati S, Ståhl FL, Kristiansson H, Solheim O, Förander P, Jakola AS. Assessment of drainage techniques for evacuation of chronic subdural hematoma: a consecutive population-based comparative cohort study. Journal of neurosurgery. 2017:1–7. doi: 10.3171/2016.12.Jns161713

17. Soleman J, Lutz K, Schaedelin S, Kamenova M, Guzman R, Mariani L, Fandino J. Subperiosteal vs Subdural Drain After Burr-Hole Drainage of Chronic Subdural Hematoma: A Randomized Clinical Trial (cSDH-Drain-Trial). Neurosurgery. 2019;85:E825–e834. doi: 10.1093/neuros/nyz095

18. Jensen TSR, Haldrup M, Hjortdal Grønhøj M, Miscov R, Larsen CC, Debrabant B, Poulsen FR, Bergholt B, Hundsholt T, Bjarkam CR, et al. National randomized clinical trial on subdural drainage time after chronic subdural hematoma evacuation. Journal of neurosurgery. 2021:1–8. doi: 10.3171/2021.10.Jns211608

19. Amoo M, O’Cearbhaill RM, McHugh P, Henry J, O’Byrne K, Ben Husien M, Javadpour M. Derivation of a Clinical Score for Prediction of Recurrence Following Evacuation of Chronic Subdural Hematoma: A Retrospective Cohort Study at a National Referral Centre. World neurosurgery. 2021;154:e743–e753. doi: 10.1016/j.wneu.2021.07.126

20. Bartley A, Bartek J, Jr., Jakola AS, Sundblom J, Fält M, Förander P, Marklund N, Tisell M. Effect of Irrigation Fluid Temperature on Recurrence in the Evacuation of Chronic Subdural Hematoma: A Randomized Clinical Trial. JAMA neurology. 2023;80:58–63. doi: 10.1001/jamaneurol.2022.4133

21. Lukasiewicz AM, Grant RA, Basques BA, Webb ML, Samuel AM, Grauer JN. Patient factors associated with 30-day morbidity, mortality, and length of stay after surgery for subdural hematoma: a study of the American College of Surgeons National Surgical Quality Improvement Program. Journal of neurosurgery. 2016;124:760–766. doi: 10.3171/2015.2.Jns142721

22. Ironside N, Nguyen C, Do Q, Ugiliweneza B, Chen CJ, Sieg EP, James RF, Ding D. Middle meningeal artery embolization for chronic subdural hematoma: a systematic review and meta-analysis. Journal of neurointerventional surgery. 2021;13:951–957. doi: 10.1136/neurintsurg-2021-017352

23. Salih M, Shutran M, Young M, Vega RA, Stippler M, Papavassiliou E, Alterman RL, Thomas A, Taussky P, Moore J, et al. Reduced recurrence of chronic subdural hematomas treated with open surgery followed by middle meningeal artery embolization compared to open surgery alone: a propensity score-matched analysis. Journal of neurosurgery. 2022:1–7. doi: 10.3171/2022.11.Jns222024

24. Nazari P, Golnari P, Ansari SA, Cantrell DR, Potts MB, Jahromi BS. Unplanned readmission after carotid stenting versus endarterectomy: analysis of the United States Nationwide Readmissions Database. Journal of neurointerventional surgery. 2022. doi: 10.1136/neurintsurg-2021-018523

